# Personalized Prescription of ACEI/ARBs for Hypertensive COVID-19 Patients

**DOI:** 10.1101/2020.10.30.20223594

**Authors:** Dimitris Bertsimas, Alison Borenstein, Luca Mingardi, Omid Nohadani, Agni Orfanoudaki, Bartolomeo Stellato, Holly Wiberg, Pankaj Sarin, Dirk J. Varelmann, Vicente Estrada, Carlos Macaya, Iván J. Núñez Gil

## Abstract

The COVID-19 pandemic has prompted an international effort to develop and repurpose medications and procedures to effectively combat the disease. Several groups have focused on the potential treatment utility of angiotensin-converting–enzyme inhibitors (ACEIs) and angiotensin-receptor blockers (ARBs) for hypertensive COVID-19 patients, with inconclusive evidence thus far. We couple electronic medical record (EMR) and registry data of 3,643 patients from Spain, Italy, Germany, Ecuador, and the US with a machine learning framework to personalize the prescription of ACEIs and ARBs to hypertensive COVID-19 patients. Our approach leverages clinical and demographic information to identify hospitalized individuals whose probability of mortality or morbidity can decrease by prescribing this class of drugs. In particular, the algorithm proposes increasing ACEI/ARBs prescriptions for patients with cardiovascular disease and decreasing prescriptions for those with low oxygen saturation at admission. We show that personalized recommendations can improve patient outcomes by 1.0% compared to the standard of care when applied to external populations. We develop an interactive interface for our algorithm, providing physicians with an actionable tool to easily assess treatment alternatives and inform clinical decisions. This work offers the first personalized recommendation system to accurately evaluate the efficacy and risks of prescribing ACEIs and ARBs to hypertensive COVID-19 patients.

**Highlights:**

− This paper introduces a data-driven approach for personalizing the prescription of ACE inhibitors (ACEIs) and angiotensin-receptor blockers (ARBs) for hypertensive COVID-19 patients.
− Leveraging an international cohort of more than 3,500 patients, we identify clinical and demographic characteristics that may affect the effectiveness of ACEIs/ARBs for COVID-19 patients, such as low oxygen saturation at admission.
− We developed a user-friendly online application that is available to physicians to facilitate interpretation and communication of the results of the algorithm.

## 1 Introduction

Since its emergence in December 2019, the COVID-19 pandemic has put an enormous strain on healthcare systems around the world. As of October 19, 2020, more than 39 million cases have been reported globally, with a death toll greater than 1.1 million [55]. Patients who have developed severe acute respiratory syndrome coronavirus 2 (SARS-CoV-2) infection exhibit a wide range of clinical responses, from being asymptomatic to being critically ill [45]. Given the heterogeneity of clinical manifestations of the disease, it is of critical importance to be able to understand how patients will respond to various potential treatments [50].

There is still limited evidence from randomized controlled trials (RCTs) to recommend specific anti-SARS-CoV-2 treatment for patients with a suspected or confirmed COVID-19 infection. A preliminary report from the RECOVERY collaborative group demonstrates that the use of dexamethasone for COVID-19 patients receiving either invasive mechanical ventilation or oxygen alone can result in lower mortality [23]. Among antivirals, remdesivir is the only drug which has shown promising results and recently received FDA approval; in a relatively small cohort of patients hospitalized for severe COVID-19, clinical improvement was observed in 68% of the participants [22]. In separate multi-center studies, it was shown that remdesivir can lead to faster clinical improvement in adults who were hospitalized with COVID-19 and had evidence of lower respiratory tract infection [3, 54]. There is controversy regarding the effects of chloroquine and hydroxychloroquine [9, 19, 20]. A wide range of other therapies are continuously being evaluated, including corticosteroids, other antiviral agents (lopinavir, ritonavir), antibodies, and convalescent plasma transfusion [56].

ACE inhibitors (ACEIs) and angiotensin-receptor blockers (ARBs) have gained attention regarding their potential benefits and harms to COVID-19 patients. ACEIs and ARBs are two medications commonly used to treat high blood pressure. They work on the same biochemical pathway in the body to treat hypertension, but at different spots. Initially, there was concern regarding a potential increased risk to COVID-19 patients taking ACEI/ARBs due to the drugs’ biological mechanisms. SARS-CoV-2 attacks human cells by binding its viral spike protein to the membrane-bound form of the monocarboxypeptidase angiotensin-converting enzyme 2 (ACE2) [27]. ACEIs and ARBs directly act on the renin angiotensin aldosterone system, raising speculation that ACE inhibitors and ARBs might be harmful in patients with the disease [17]. However, multiple clinical investigations from various countries showed that neither ACEIs nor ARBs were associated with an increased risk of in-hospital death or severe COVID-19 [31, 34, 37, 39, 43]. To the contrary, among hospitalized patients with COVID-19 and coexisting hypertension, inpatient use of ACEI/ARBs was associated with lower risk of all-cause mortality [58]. The effects of ACEI/ARBs for hypertensive COVID-19 patients are therefore not well-understood, and there is no consensus on appropriate uses of these drugs [53].

Personalized or precision medicine aims at providing answers to these types of questions [24]. This emerging field is expected to radically transform medical care and public health, uncovering prevention and treatment programs more closely targeted to the individual patient [29]. Machine learning (ML) and analytics play a major role in this endeavor [15]. By leveraging large datasets, these techniques can generate insights and derive decision rules by processing information that exceed the capacity of the human brain [42]. Thus, they are able to exploit data patterns at the individual level to determine the effect of a treatment or the projected risk of mortality/morbidity.

### 1.1 Literature Review

Our objective is to develop a model that determines whether a treatment *T* can reduce the risk of mortality/morbidity for an individual patient. We include in our dataset *n* observations of the 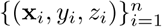, where **x**_*i*_ ∈ ℝ ^*p*^ are the features of the *i*th observation, *z*_*i*_ ∈ [*T, C*] is the assigned treatment or control, and *y*_*i*_ ∈ ℝ is the corresponding outcome of interest. We let *y*(*C*) be the potential outcome resulting from the assignment of the control and *y*(*T*) of the treatment.

This problem lies at the core of the causal inference literature. Rubin [47] set its foundation by proposing the Potential Outcomes Framework which assumes that patients are prescribed a treatment via a probabilistic assignment mechanism. Under this framework, the causal effect of a treatment *T* is measured by the difference in the potential outcomes *y*(*T*) − *y*(*C*). The fundamental challenge of this problem is that for any given patient, only one of the potential outcomes is observed [1, 48]. As a result, causal inference methodologies usually focus on estimating the aggregated treatment effect, studying its impact on an entire population rather than at the individual level.

Personalized medicine calls for more individualized approaches that leverage patient-level characteristics to evaluate treatment efficacy for each patient in isolation. Since machine learning estimates a binary or continuous outcome of interest from large, high-dimensional datasets, a common approach involves training separate prediction models for the treatment and the control group, and recommending the alternative with the best outcome [18, 41]. This technique is referred in the literature as “Regress and Compare” [51]. Bertsimas et al. [5] showed how this framework can be extended for the management of diabetes by applying the *k*-nearest neighbors method. While a useful and intuitive framework, “Regress and Compare” has received criticism as it can be subject to prediction errors and biases associated with the specific classification or regression algorithm. More advanced machine learning approaches are focused on tree-based methods that involve recursive partitioning [30], causal trees [2], causal forests [52], and optimal prescriptive trees [6].

Recently, a machine learning based framework was proposed to identify the best therapy for patients with coronary artery disease [8]. The authors created a series of regression models for several treatment alternatives to predict the time from diagnosis to a potential heart attack or a stroke. It extends the classical “Regress and Compare” approach by aggregating an ensemble of ML models, making it more robust to individual method biases. The algorithm recommends the therapy with the best expected outcome through a voting mechanism that considers the predictions from each of the regression models. We build upon this framework and adapt it to the specific challenges posed by COVID-19.

### 1.2 Contributions

In this paper, we propose a machine learning-based approach for personalized prescription of ACEI/ARBs for hospitalized hypertensive patients with COVID-19. We leverage EMR and registry data of 3,643 patients from Spain, Italy, Germany, Ecuador, and the US to provide accurate predictions of expected mortality and morbidity. We then propose individualized treatment decisions by applying the voting scheme that was introduced by Bertsimas et al. [8]; we combine multiple binary classification models to identify whether there is a potential benefit from prescribing this class of drugs based on a patient’s characteristics. The main contributions of this work can be summarized as follows:

− We combine EMR data with an international registry to create a diverse dataset from multiple clinical centers. We present a unified dataset from 38 hospitals of five distinct countries, encompassing demographics, pre-admission comorbidities and medications, vitals at admission, laboratory test results, and inpatient medications.
− We develop binary classification models to predict mortality and morbidity during hospital admission under treatment alternatives.
− We utilize an ensemble analytical framework, that has been previously applied to personalize treatments for coronary artery disease and hypertension [7, 8], to evaluate the effectiveness of ACEI/ARBs at the individual level.
− We discover specific patient populations who benefit most from this class of drugs, such as patients with cardiovascular disease, as well as those who may suffer from these prescriptions, like patients with low oxygen saturation at admission. We provide clinical insights that validate findings from the medical literature, and propose new hypotheses for further investigation.
− We provide a dynamic online application with a user-friendly interface of the predictive models and the resulting prescriptions for use by clinical providers.

## 2 Methods

We propose a machine learning approach to the problem of personalizing treatments. A patient’s prescription is generated based on individualized risk scores under each treatment alternative. We leverage clinical data from 3,643 patients across international institutions to train our models. One ensemble of various machine learning models is trained to predict mortality/morbidity risk with ACEI/ARBs, and another ensemble is trained to predict the risk when patients are not given ACEI/ARBs. We then employ a voting scheme to aggregate the risk scores of the individual methods and give a final prescription and estimated benefit of treatment. An overview of the approach is illustrated in Figure 1.

**Fig 1:**
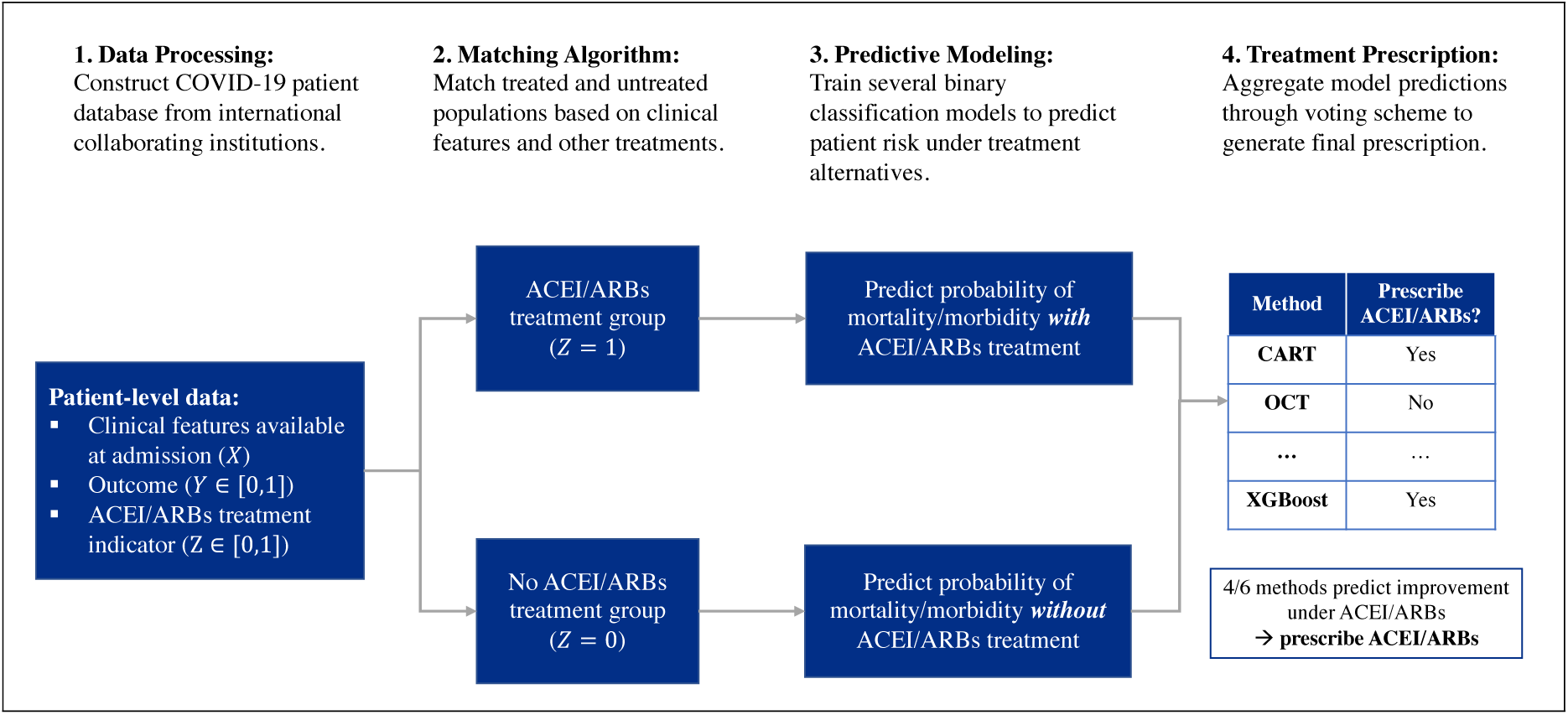
An overview of the machine learning approach to prescription personalization.

### 2.1 Data Resources

This study utilizes patient data from 38 hospitals across five countries: Spain, Italy, Ecuador, Germany, and the United States. Depending on the institution, data is sourced either from a standardized COVID-19 specific registry or from electronic medical records. The data is separated into a derivation cohort, which is used to train the machine learning models, and a validation cohort, which is used to test the models on unseen populations. The derivation cohort is comprised of data from 2,842 hypertensive patients from HOPE registry’s hospitals in Spain and from HM Hospitals, also in Spain [21]. The validation cohort consists of data from 801 patients diagnosed with hypertension from the following organizations and geographic locations: HOPE (Italy, Germany, Ecuador), ASST Cremona (Northern Italy), and Brigham and Women’s Hospital (Massachusetts, United States). The study population includes adult patients with a hypertension diagnosis who were admitted to the hospital with confirmed severe acute respiratory syndrome coronavirus 2 (SARS-CoV-2) infection by polymerase chain reaction testing of nasopharyngeal samples. Hypertension was identified using diagnosis codes from a patient’s medical record or from patient history available in the registry, as accepted in their respective medical centers or attending medical teams; this is detailed further in the Appendix. A description and details of the collaborating organizations, as well as the time horizon of admissions for each organization’s study population, can be found in Table 1.

**Table 1:** Overview of participating institutions in the study. The column NH stands for Number of Hospitals.

| Organization | Region | Study Dates | NH | Description |
| --- | --- | --- | --- | --- |
| **Derivation Cohort** |  |  |  |  |
| **HOPE (Spain)** | Madrid, Galicia, Castilla y León, Castilla la Mancha, Andalucía, Murcia, Valencia, Cataluña (Spain) | 03/01-04/30 | 21 | HOPE is an international registry that was created at the beginning of the pandemic with the aim of collecting data to carefully characterize the clinical profile of patients infected with COVID-19. The study was initiated by the Hospital Clinico San Carlos in Madrid and the majority of the recorded patients were hospitalized in Spain. |
| **HM Hospitals** | Madrid, Galicia, Castilla y León, Cataluña (Spain) | 02/01-04/20 | 17 | HM Hospitals, a leading Hospital Group in Spain with 15 general hospitals and 21 clinical centres that cover the regions of Madrid, Galicia, and León. The group has served more than 2,300 COVID-19 patients over the last two months. Its total capacity includes more than 1,468 beds and 101 operating rooms. |
| **Validation Cohort** |  |  |  |  |
| **ASST Cremona** | Lombardy (Italy) | 02/01-05/08 | 3 | Azienda Socio-Sanitaria Territoriale di Cremona (ASST Cremona) includes the Ospedale di Cremona, Ospedale Oglio Po and other minor public hospitals in the Province of Cremona. Cremona is one of the most hit Italian provinces in Lombardy in the Italian COVID-19 crisis with a total of 4,422 positive cases to date. Ospedale di Cremona has around 750 beds. During the COVID-19 crisis all elective activities and surgeries were suspended and most of the hospital was converted to treat COVID-19. |
| **HOPE (Other)** | Mannheim (Germany), Lombardy, Piedmont, Lazio, Puglia, Marche (Italy), Guayaquil, Quito (Ecuador) | 03/01-04/30 | 14 | This subpopulation includes patients discharged (deceased or alive) from all collaborating hospital centers from the HOPE registry outside of Spain with a confirmed diagnosis or a COVID-19 high suspicion have been included. There are no exclusion criteria, except for the patient's explicit refusal to participate. |
| **Brigham and Women's Hospital** | Massachusetts (USA) | 03/01-05/31 | 1 | Brigham and Women's Hospital is a leading academic medical center located in Boston, MA. Today it is part of Massachusetts General Brigham (MGB), which comprises 16 institutions in New England. During the COVID-19 pandemic, it has played a central role providing health care services and conducting research with multiple academic institutions of the US. |

### 2.2 Clinical Features

Dataset features include demographics, pre-admission comorbidities and medications, vitals at admissions, laboratory test results, and inpatient medications. In total, we compile 29 features, which are summarized in Supplemental Tables 3-4. Comorbidities are derived from the International Classification of Diseases (ICD), 9th and 10th revision, using codes of hospital discharges. Medications are extracted from the Anatomical Therapeutic Chemical (ATC) Classification System. We record the earliest laboratory test results obtained within the hospital admission and include both binary measurements (e.g., D-dimer *≥* 0.5mg/L) and continuous measurements (e.g., creatinine in mg/dL). We also collect information on patient mortality, as well as inpatient development of specific morbidities during hospitalization, including: sepsis, acute renal failure, heart failure, and embolic event. The outcome of interest is the occurrence of mortality or morbidity during hospital admission. Missing values are imputed using multivariate imputation by chained equations (MICE) (details can be found in the Appendix) [12]. We exclude all features that are not present for at least 70% of the observations.

**Table 2:** Descriptive summary of clinical characteristics of derivation and validation populations prior to matching.

|  | Derivation |  | Validation |  |
| --- | --- | --- | --- | --- |
|  | ACEI/ARBs | No ACEI/ARBs | ACEI/ARBs | No ACEI/ARBs |
| Patient Count | 1043 | 1663 | 280 | 521 |
| Age | 70.0 (57.0-79.0) | 70.0 (56.5-79.0) | 68.0 (56.0-78.0) | 68.0 (57.0-79.0) |
| Gender = Male | 599 (57.4%) | 985 (59.2%) | 175 (62.5%) | 336 (64.5%) |
| Race = Black | 2 (0.2%) | 1 (0.1%) | 8 (2.9%) | 40 (7.7%) |
| Race = Caucasian | 955 (91.6%) | 1532 (92.1%) | 237 (84.6%) | 369 (70.8%) |
| Race = Hispanic | 78 (7.5%) | 109 (6.6%) | 34 (12.1%) | 106 (20.3%) |
| Race = Asian | 3 (0.3%) | 6 (0.4%) | 1 (0.4%) | 5 (1.0%) |
| Temperature | 36.8 (36.5-37.5) | 36.8 (36.6-37.4) | 37.3 (36.8-37.8) | 37.0 (36.8-37.8) |
| Creatinine (mg/dL) | 0.9 (0.7-1.2) | 0.9 (0.7-1.2) | 1.0 (0.8-1.3) | 1.0 (0.8-1.3) |
| Sodium (mmol/L) | 138.0 (135.0-140.0) | 138.0 (135.0-140.0) | 138.0 (135.0-140.0) | 138.0 (135.0-140.0) |
| Hemoglobin (g/dL) | 14.0 (12.5-15.0) | 14.0 (12.9-15.0) | 13.6 (12.0-14.8) | 13.0 (11.4-14.2) |
| Leukocytes (1e3/μL) | 6.4 (4.9-8.5) | 6.3 (4.8-8.4) | 6.9 (5.0-9.8) | 7.0 (5.1-9.4) |
| Lymphocytes (1e3/μL) | 1.0 (0.7-1.4) | 1.0 (0.7-1.4) | 1.0 (0.7-1.4) | 1.0 (0.8-1.4) |
| Platelets (1e3/μL) | 194.0 (150.0-255.0) | 195.0 (152.0-250.0) | 200.5 (150.8-256.2) | 204.0 (152.0-285.0) |
| Low Oxygen Saturation | 345 (33.1%) | 614 (36.9%) | 114 (40.7%) | 182 (34.9%) |
| Low Systolic BP | 78 (7.5%) | 191 (11.5%) | 10 (3.6%) | 35 (6.7%) |
| Elevated D-Dimer | 728 (69.8%) | 1244 (74.8%) | 224 (80.0%) | 419 (80.4%) |
| Elevated CRP | 945 (90.6%) | 1564 (94.0%) | 193 (68.9%) | 429 (82.3%) |
| Elevated Transaminases | 406 (38.9%) | 688 (41.4%) | 125 (44.6%) | 239 (45.9%) |
| Elevated LDH | 758 (72.7%) | 1298 (78.1%) | 124 (44.3%) | 227 (43.6%) |
| Diabetes | 315 (30.2%) | 497 (29.9%) | 59 (21.1%) | 159 (30.5%) |
| Hypertension | 1043 (100.0%) | 1663 (100.0%) | 280 (100.0%) | 521 (100.0%) |
| Dislipidemia | 517 (49.6%) | 823 (49.5%) | 72 (25.7%) | 199 (38.2%) |
| Obesity | 310 (29.7%) | 458 (27.5%) | 54 (19.3%) | 129 (24.8%) |
| Renal Insufficiency | 84 (8.1%) | 192 (11.5%) | 18 (6.4%) | 65 (12.5%) |
| Lung Disease | 285 (27.3%) | 430 (25.9%) | 45 (16.1%) | 122 (23.4%) |
| Atrial Fibrillation | 70 (6.7%) | 161 (9.7%) | 13 (4.6%) | 47 (9.0%) |
| HIV | 2 (0.2%) | 4 (0.2%) | 0 (0.0%) | 1 (0.2%) |
| Heart Disease | 360 (34.5%) | 603 (36.3%) | 71 (25.4%) | 154 (29.6%) |
| Cerebrovascular Disease | 96 (9.2%) | 190 (11.4%) | 14 (5.0%) | 56 (10.7%) |
| Connective Tissue Disease | 52 (5.0%) | 72 (4.3%) | 8 (2.9%) | 44 (8.4%) |
| Liver Disease | 66 (6.3%) | 75 (4.5%) | 5 (1.8%) | 29 (5.6%) |
| Cancer | 155 (14.9%) | 310 (18.6%) | 19 (6.8%) | 77 (14.8%) |
| Corticosteroids | 387 (38.0%) | 724 (44.1%) | 138 (49.3%) | 225 (44.3%) |
| Interferons | 108 (10.6%) | 176 (10.8%) | 50 (17.9%) | 47 (9.3%) |
| Tocilizumab | 96 (9.2%) | 176 (10.6%) | 20 (7.1%) | 20 (3.8%) |
| Antibiotics | 842 (80.7%) | 1367 (82.2%) | 221 (78.9%) | 399 (76.6%) |
| Death | 219 (21.0%) | 352 (21.2%) | 67 (23.9%) | 139 (26.7%) |
| Mortality/Morbidity | 329 (31.5%) | 545 (32.8%) | 82 (29.3%) | 179 (34.4%) |
| Heart Failure | 73 (7.0%) | 92 (5.5%) | 12 (4.3%) | 22 (4.2%) |
| Acute Renal Failure | 163 (15.6%) | 266 (16.0%) | 19 (6.8%) | 43 (8.3%) |
| Sepsis | 108 (10.4%) | 148 (8.9%) | 8 (2.9%) | 26 (5.0%) |
| Embolic Event | 14 (1.3%) | 25 (1.5%) | 4 (1.4%) | 7 (1.3%) |
| Mechanical Ventilation | 53 (5.1%) | 82 (4.9%) | 23 (8.8%) | 44 (12.1%) |

**Table 3:** AUC of the six binary classification algorithms trained on two populations. The twelve models are evaluated on the training, testing, and validation datasets.

|  | Training Data |  | Testing Data |  | Validation Data |  |
| --- | --- | --- | --- | --- | --- | --- |
|  | ACEI/ARBs | No ACEI/ARBs | ACEI/ARBs | No ACEI/ARBs | ACEI/ARBs | No ACEI/ARBs |
| **RF** | 0.886 | 0.862 | 0.834 | 0.814 | 0.770 | 0.761 |
| **CART** | 0.843 | 0.827 | 0.812 | 0.790 | 0.762 | 0.714 |
| **OCT** | 0.879 | 0.829 | 0.797 | 0.777 | 0.737 | 0.699 |
| **XGBOOST** | 0.909 | 0.927 | 0.819 | 0.802 | 0.768 | 0.743 |
| **QDA** | 0.883 | 0.870 | 0.827 | 0.813 | 0.718 | 0.736 |
| **GB** | 0.824 | 0.813 | 0.826 | 0.805 | 0.710 | 0.734 |
| **Average AUC** | 0.871 | 0.855 | 0.819 | 0.800 | 0.744 | 0.731 |

### 2.3 Covariate Matching

As opposed to data obtained from a randomized controlled trial (RCT), for which treatment assignment is random, the data in our study is observational in nature. Given that we aim to determine the effect of a treatment on the probability of mortality and morbidity for a patient, we must consider that individuals taking a specific treatment may differ from those that are not taking the treatment in terms of their baseline, or pre-treatment, characteristics. Such characteristics may affect both treatment assignment and mortality/morbidity risk and may, therefore, confound our treatment effect estimates. We also recognize that the medications that we are investigating were not used in isolation and were often administered in combination with other medications; this, too, may bias our treatment effect estimates. In order to mitigate bias introduced by confounding variables, we use matching techniques prior to training our machine learning models.

Matching is a method that can be used to control for confounding in observational studies. The motivation of the technique is to find groups of treated and non-treated individuals whose pre-treatment characteristics are similar, and to then create models using only these individuals. By minimizing the differences in pretreatment characteristics, we become more confident that the outcome estimates can be attributed to differences in the treatment assignment rather than to preexisting differences between the individuals in the treated and non-treated group. We evaluate our matching procedure based on how balanced pre-specified covariates are between our treated and non-treated groups. Balance is measured by comparing the pairwise absolute standardized mean differences in covariates. Groups are considered well-balanced if their standardized mean differences are below 0.10.

For our matching procedure, we aim to find populations of patients in each treatment group that (1) have similar baseline characteristics and (2) have similar medications as part of their additional treatments. To study of the effect of ACEI/ARBs on patient mortality, we first identify all of the patients that were given either ACEIs or ARBs during their hospital stay. We then use cardinality matching [59] to identify the most similar cohort of non-ACEI/ARB recipients to this group, as measured by a set of important pre-treatment and other treatment features. The final dataset we use to predict patient mortality consists of the ACEI/ARBs treatment group along with the matched dataset from the non-ACEI/ARBs treatment patient group.

### 2.4 Risk Prediction under Treatment Alternatives

Using the matched datasets, two sets of models are constructed to predict a patient’s risk of mortality/morbidity, as defined in Section 2.2. One set of models is trained on patients who were given ACEI/ARBs, and the other on patients who were not given ACEI/ARBs. The matching process aims to equalize the baseline characteristics of the populations to better isolate the effect of ACEI/ARBs between the two sets of models.

For each treatment, we train six binary classification models to predict a patient’s risk of mortality/morbidity. The machine learning methods we utilize are: random forests [10], classification and regression trees [11], optimal classification trees [4], gradient boosted decision trees [14], quadratic discriminant analysis [25], and Gaussian naïve Bayes [57]. These models take diverse approaches to classification tasks and involve tradeoffs in their interpretability, handling of nonlinear relationships, and computational complexity. Further details on the training procedures and parameter tuning employed for these models are available in the Appendix. Algorithms were trained using Python 3.6.3 and Julia 1.2.0 through Scikit-learn [40], XGBoost [14], and the Interpretable AI [28] packages.

The primary metric of performance for the classification models is Area Under the ROC Curve (AUC), which measures a model’s ability to discriminate between high and low risk patients. Although the predictive models are not the final output of our framework, this evaluation is important to verify that the individual models provide high quality predictions.

We apply the SHapley Additive exPlanations (SHAP) to identify the most important risk drivers for each learner under both treatment alternatives [35, 36]. We use the SHAP Python package [36], leveraging the Tree Explainer for the XGBoost, classification and regression trees, and random forests algorithms and the Kernel Explainer for the logistic regression, quadratic discriminant analysis, and Gaussian Naive Bayes classifiers. The SHAP methodology approximates any nonlinear prediction model with a linear model around the patient prediction. The coefficients of the linear approximation are called SHAP values. They are computed for each observation by introducing every feature separately and comparing the model output risk. We calculate the absolute mean SHAP value for all the independent covariates using the testing set. We report the ones with the greatest impact on the prediction task.

### 2.5 Treatment Prescription Methodology

Each algorithm is used to train two separate models: one model with ACEI/ARBs and one without ACEI/ARBs. For a given patient and algorithm, the models yield a prediction for the patients mortality/morbidity risk with ACEI/ARBs, *ŷ*_*Y*_, and without ACEI/ARBs, *ŷ*_*N*_. The algorithm recommends ACEI/ARBs if administering the treatment is predicted to have a reduction of at least 5% in the probability of mortality/morbidity. Namely, treatment with ACEI/ARBs is suggested if:

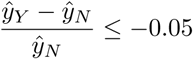

The improvement threshold is intended to reduce unnecessary prescriptions: if the patient’s predicted risk is nearly identical under both treatment alternatives, we do not recommend treatment. The threshold of 5% was chosen based on tradeoffs between treatment effectiveness and number of prescriptions; this is further explored in the Appendix.

The six ML algorithms yield six “votes” for whether or not to recommend ACEI/ARBs. The final prescription aggregates the votes. If there is majority consensus (i.e. if at least four methods agree on the optimal treatment), the majority choice is selected. In the case of ties (i.e. three methods vote for ACEI/ARBs and the other three vote against it), we consider the AUC of the individual methods as a way of measuring the credibility of the votes. We select the treatment option which has a higher average AUC for the methods that voted for it. In other words, in the case of a tie between the treatments, we follow the treatment selected by the most credible methods.

### 2.6 Prescription Evaluation

An effective treatment prescription scheme should improve outcomes compared to the current standard of practice. Since our outcome of interest is a patient’s mortality/morbidity, good prescriptions would *decrease* the incidence rate. Prescription evaluation is a well-recognized problem due to the lack of counterfactuals; we only have data on the treatment received by a patient, and we cannot know what their outcome would have been under the other treatment option. We must therefore estimate the counterfactuals to evaluate the quality of our prescriptions, and we can leverage the predictive models for this task. We perform this assessment in several ways:

#### Prescription Effectiveness

The effectiveness of the prescription scheme can be estimated by comparing the actual event rate to the average predicted risk under our prescription scheme. For a given patient, we can compute their predicted risk under a treatment as the average probability among the methods that voted for the treatment; for example, if ACEI/ARBs are selected by four methods, the predicted probability with treatment would be the averages of these four methods’ predictions. Let *ŷ*_*ip*_ denote patient *i*’s predicted probability of mortality/morbidity under the recommended treatment, and *y*_*i*_ ∈ {0, 1} indicate the true outcome. Then the prescription effectiveness (PE) is defined as:

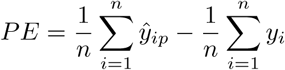

If the raw mortality/morbidity rate is 30%, for example, and the average probability of mortality/morbidity is 25% under the prescription scheme, then the PE equals -0.05. We adjust the calculation of this metric to include only cases for which the algorithmic recommendation differs to the doctors’ prescription at the standard of care. Thus, observations of patients whose medication did not change were not included. Note that a negative number indicates an improvement in mortality/morbidity.

#### Calibrated Prescription Effectiveness

When applying the prescription algorithm to external populations with significantly different mortality/morbidity rates, the PE metric may require recalibration. PE compares the baseline mortality/morbidity rate to the average probability of the proposed treatments. If a new population has a much higher event rate than the training population, the predicted probabilities may be systematically low; the opposite is true if the new population has a much lower incidence rate. We take a simple rescaling approach to adjust the probabilities proportionally to the incidence rates. Denoting the outcome rate on the training population as 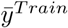, we can construct a calibration factor

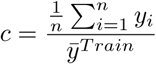

If the new population has a higher incidence rate, this factor will be greater than 1, meaning that we scale up the projected probabilities. If the new population has a lower incidence rate, then *c <* 1 and the probabilities will be scaled down. The calibrated PE (CPE) is then given by:

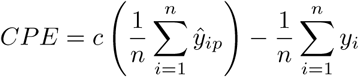

Other sophisticated calibration schemes exist, but this metric has an appeal of not requiring access to data and outcomes for the full external population. For example, if we are applying the method to a new hospital, we only need to know the baseline mortality/morbidity to recalibrate our probabilities. We also note that by using a constant scaling factor, we preserve the ordering of the probabilities; this, therefore, does not affect the prescription decisions or the AUC of the models.

#### Prescription Robustness

While PE and CPE are highly intuitive metrics, they can also be biased in their estimates of the outcome probability under the prescription scheme since the predictions are taken from methods that also determine the prescription. In the binary classification setting, these metrics involve data of different types as they compare the discrete outcomes with continuous probabilities. Prescription robustness (PR) takes a more objective view: it uses a single ML method to evaluate both the outcome probability under the standard of care (given treatments) and the probability under the prescription. For example, PR with respect to CART would be computed as:

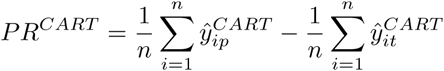

where 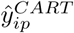and 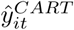 are the CART models’ predicted outcomes under the proposed treatment (*p*) and true treatment (*t*), respectively. PR is calculated for each of the six candidate methods, and the range across the methods is reported.

#### Treatment Agreement Rate

We report the proportion of our prescriptions that match the true treatment decisions. For example, if out of 100 patients, 60 of our treatment recommendations are consistent with the treatment decision, then the agreement rate would be 60%. We note however that given the rapid evolution of the pandemic and shifting treatment protocols, the true treatments do not necessarily reflect a consistent treatment strategy. While the agreement rate is informative, the goal of the prescriptive algorithm is to improve upon the current standard of care and thus a high agreement rate is not necessarily desirable.

#### Prescription AUC

We also report the AUC of the risk probabilities for our prescriptions compared to the true outcomes. We can only compare the probabilities and outcomes for the patients whose treatment prescriptions *agree* with their true treatments, since we do not have the counterfactual outcomes. For example, if the agreement rate is 60%, the AUC of our prescriptions can only be compared for the 60% of patients with agreement. This metric does not assess the prescription benefit, but rather the quality of our predictions under the prescription algorithm.

## 3 Results

In this section, we present the results of our analysis from the predictive and prescriptive components of this study. In Section 3.1, we provide information regarding the final dataset and describe the impact of the matching process. Section 3.2 focuses on the predictive performance of the binary classification models trained to predict mortality/morbidity risk. Section 3.3 outlines the quantitative results of the proposed prescription mechanism. Section 3.4 summarizes the online interface that was developed to communicate the output of the algorithm to the clinical audience.

### 3.1 Data Processing

The derivation cohort contains 1,043 observations of patients receiving ACEI/ARBs and 1,663 records that are not prescribed the specific class of drugs.

Following the method described in Section 2.3, we identify optimal matches for the treatment group and restrict our cohort to an equally balanced set of 1,920 cases. We select the features for matching through a *t−*hypothesis test, identifying the variables that are most significant in differentiating those who experienced the outcome of interest and those who did not. Thus, we selected 21 patient features and also included all available covariates related to other administered treatments, including hydroxychloroquine, antivirals (lopinavir and ritonavir), corticosteroids, anticoagulants, and interferons. We achieve pairwise balance between two groups below 0.05 for all covariates considered. Figure 2 provides an illustration of the matching results. In Supplementary Table S5, we summarize the pre-treatment variables before and after the matching procedure.

**Fig 2:**
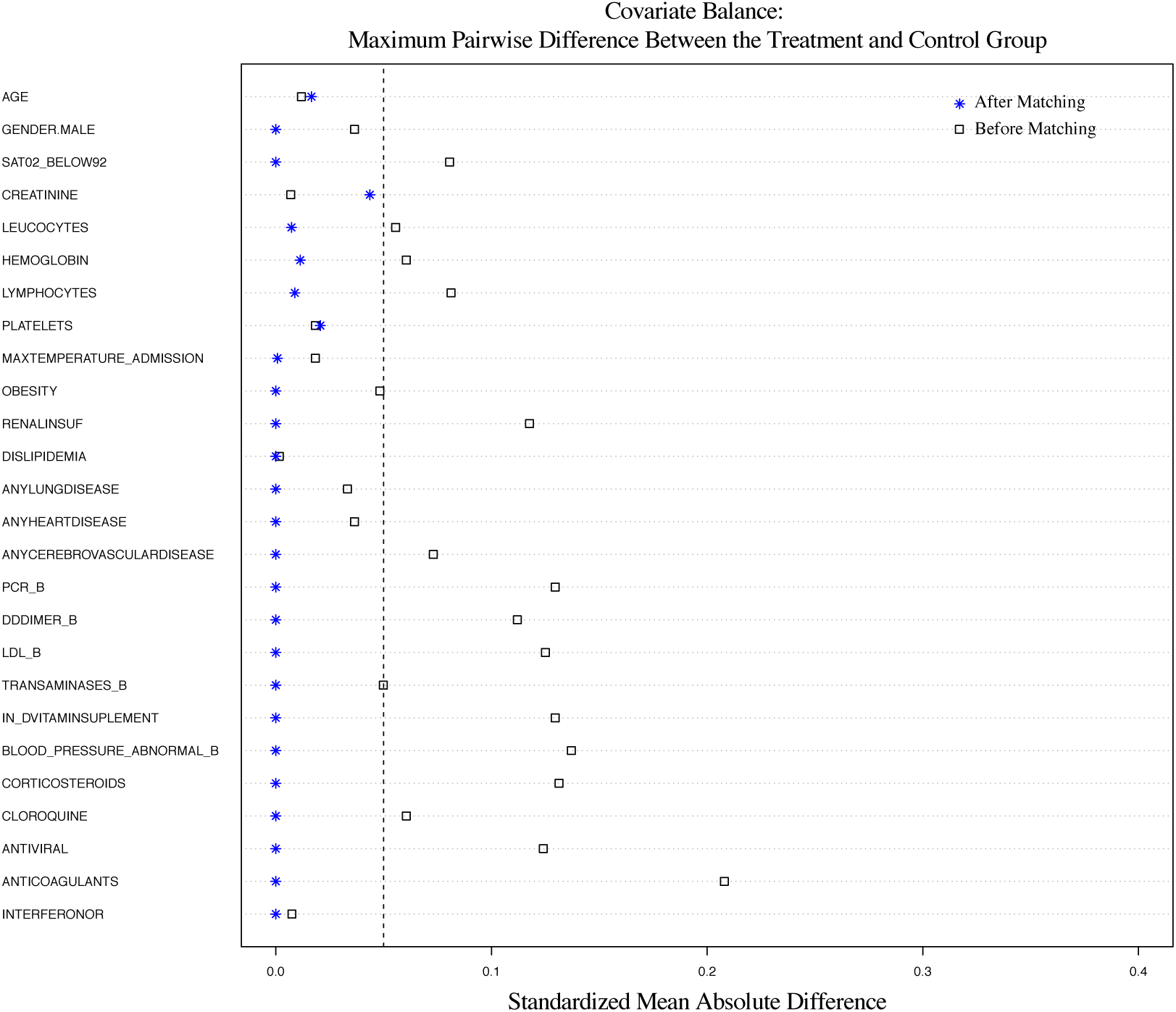
Pre-Treatment covariate balance after matching.

A descriptive summary of each treatment group’s clinical features and outcomes, for both the derivation and validation groups, is shown in Table 2. After matching, the derivation cohort has an even split of patients with and without ACEI/ARBs. The validation cohort has 801 total patients, of which 280 (35.0%) receive ACEI/ARBs.

### 3.2 Predictive Models

The AUCs for the six individual binary classification algorithms for both the ACEI/ARBs and non-ACEI/ARBs models are shown in Table 3. We report the average AUCs on the training and testing splits of the derivation population, as well as the external validation population. In general, the models for predicting outcomes without ACEI/ARBs have higher performance on the test and validation set than those for patients treated with ACEI/ARBs. Random Forests and XGBoost are the highest performing methods overall, although nearly all methods demonstrate AUCs above 0.7 across all cohorts, and above 0.8 in most cases.

A summary of the predictive models and their feature importance is shown in Table 4. Our analysis reveals that the key predictors of mortality and morbidity are common between the two treatment groups. Abnormal creatinine levels, white blood cell count, and hemoglobin are identified in both groups as the most significant lab values. Age and low oxygen saturation are also identified within the top five predictors of risk. These biomarkers have been identified in other retrospective analysis of mortality outcomes of COVID-19 [13, 32, 46]. In accordance with the medical literature [33], lymphocyte count is also found as an important feature in both models. There are a few less significant variables in each cohort that are distinct between the two groups. Platelets were found to only be a significant risk predictor for ACEI/ARBs, while blood sodium and temperature only appeared in the No ACE/ARBs cohort.

**Table 4:**
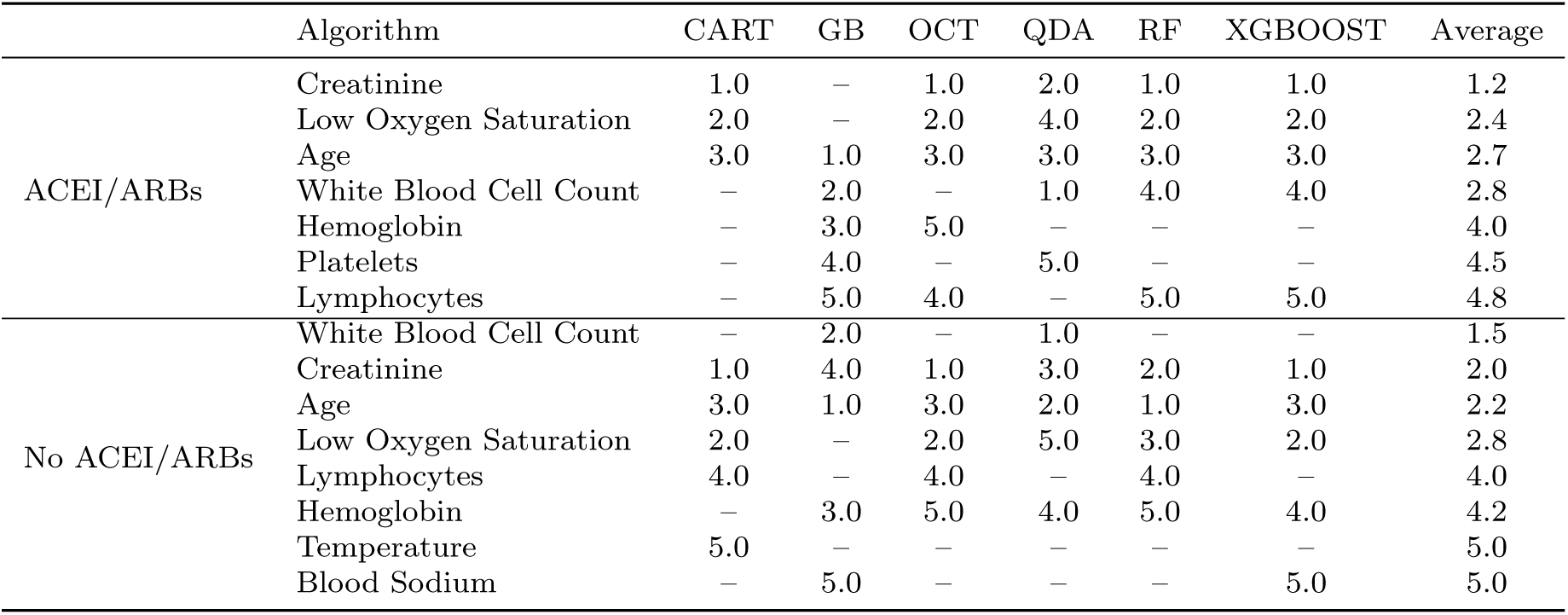
Summary of variable importance for each model by rank (1 = most important).

### 3.3 Treatment Prescriptions

Table 5 shows the results of the prescription voting scheme on the training, testing, and validation populations for an improvement threshold of 5.0%. 42.2% of the patients in the training population are recommended to receive ACEI/ARBs under our scheme, as well as 43.1% of the testing population and 46.8% of the validation population. The voting behaviors of the individual methods are included in the Appendix. The proposed prescription rate decreases from the 50% seen in practice in the training and testing data due to the matching procedure. In the raw validation data, we see an increase in the prescription rate from the observed 35.0% of COVID-19 patients who received ACEI/ARBs at these sites.

**Table 5:** Summary of prescription results on training, testing, and validation datasets, using a 5% improvement threshold.

|  | Match Rate | Presc. Count | Avg. AUC | PE | CPE | PR (Low) | PR (High) |
| --- | --- | --- | --- | --- | --- | --- | --- |
| **Training Data** | 0.521 | 688 | 0.896 | -0.058 | -0.058 | -0.008 | -0.034 |
| **Testing Data** | 0.514 | 124 | 0.804 | 0.008 | 0.007 | -0.008 | -0.033 |
| **Validation Data** | 0.482 | 375 | 0.774 | -0.010 | -0.005 | -0.007 | -0.045 |

The PE metric indicates a reduction in mortality and morbidity rate of 1.0% on the validation set, a notable reduction from the baseline mortality/morbidity rates seen in practice. When calibrating for the incidence rates in the testing and validation populations, the CPE decreases from the PE but still demonstrates a reduction in average risk. The PE metric indicates a slight increase in the mortality/morbidity probability on the test set (−0.8%).

When evaluating the given vs. recommended treatments using each of the individual ML algorithms as the ground truth, we see a benefit ranging from 0.8% to 3.3% on the test set and 0.7% to 4.5% on the validation set. Thus, we see a benefit even in the most pessimistic estimates. We observe that the PE metric can be worse than even the most pessimistic PR estimate; this is due to the fact that PE compares probabilities to the event rate, versus directly comparing probabilities. While both are useful, PR provides a more consistent basis for evaluation.

The match rate is 51.4% on the testing set and 48.2% on the validation set. The predictions of mortality/morbidity under our prescriptions have an AUC of 80.4% on the testing set and 77.4% on the validation set, demonstrating strong discriminative ability for risk in cases where the predictions match the true treatment decisions.

Figure 3 compares the proposed prescription frequencies to the true treatment decisions administered in practice on the validation data, broken down by various clinical features. These figures offer insight into how our prescriptions differ from current practice and how the treatment patterns change for specific clinical characteristics.

**Fig 3:**
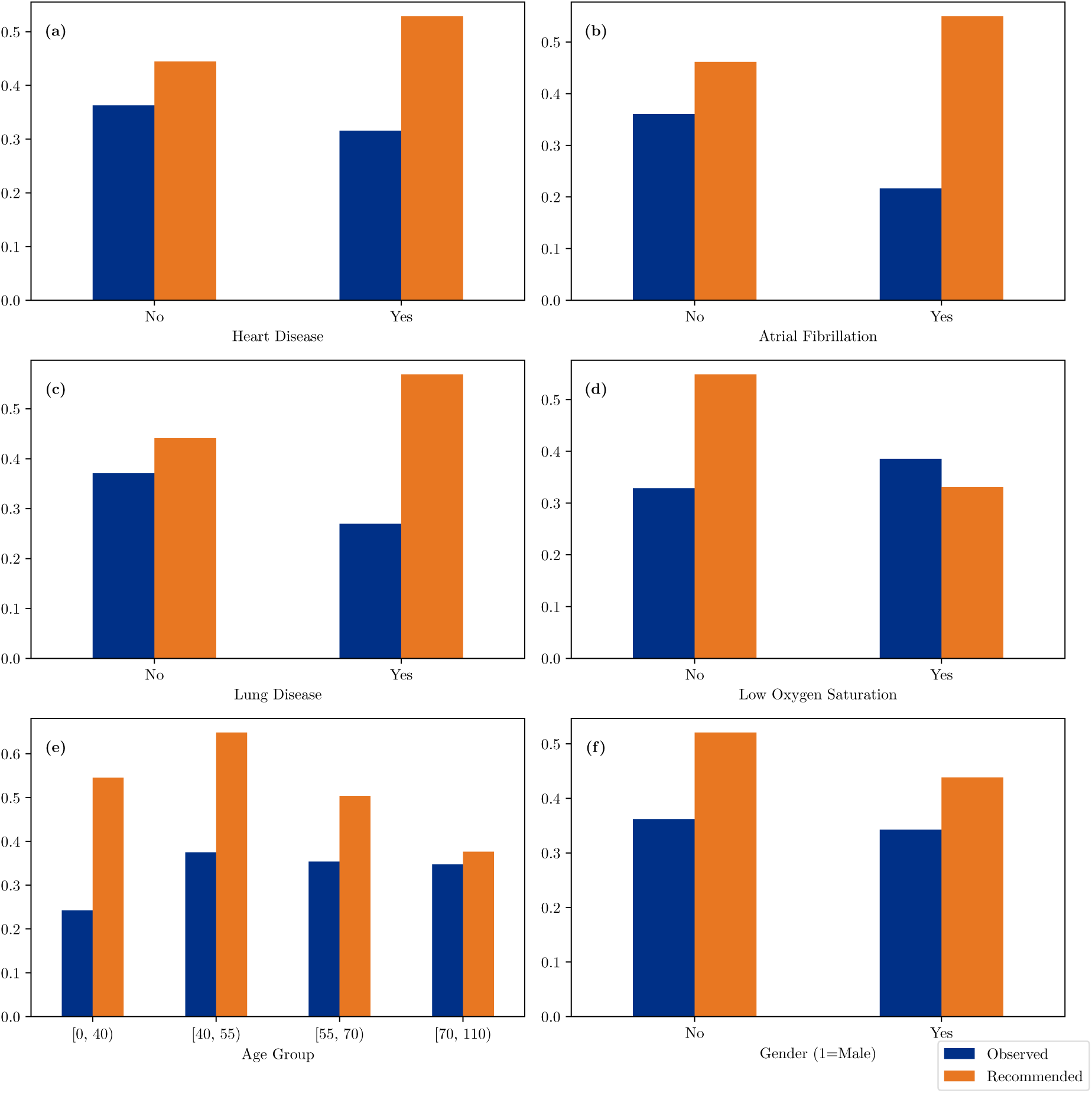
Comparison of ACEI/ARBs prescription rates under observed (blue) and recommended (orange) treatments for the validation dataset.

Our results generally propose to increase the prescription rate in the validation data, which treated 35.0% of patients with ACEI/ARBs in practice. The prescription rate increases by 67.6% for patients with heart disease and 22.5% for patients without the comorbidity. Specifically, for the case of atrial fibrillation, our algorithm suggests a significant increase by 153.8% for those diagnosed with the disease, compared to 28.1% for those who are not. Notice that for patients with chronic lung disease, the proposed prescription is 237.5% higher compared to the standard of care. The corresponding increase for patients who were not diagnosed with lung disease is lower, 19.1%. The personalized approach leads to a decrease in the prescription rate by -14.0% for patients who were admitted with low oxygen saturation. To the contrary, observations with oxygen saturation levels at the normal range were associated with a 66.9% increase. The prescription rate increases across all age groups, with greater increases for younger patients; the algorithm raises prescriptions by 125.0% for patients below 40 and only 8.3% for patients over 70. Finally, the algorithm proposes to increase the prescription rate more for women (43.8%) than for men (28.0%).

### 3.4 Online Algorithm Interface

Our goal is to provide clinicians with a readily available and actionable tool that can communicate the algorithm recommendations. For this reason, we have developed an online application that can directly inform the decision making process of physicians using the proposed models. Through this application (accessible at: https://www.covidanalytics.io/treatments), practitioners can enter new patient data at hospital admission, obtain individualized estimations of mortality/morbidity risk and evaluate the effectiveness of ACEI/ARBs for their own patients. Figure 4 displays the user interface of the web application.

**Fig 4:**
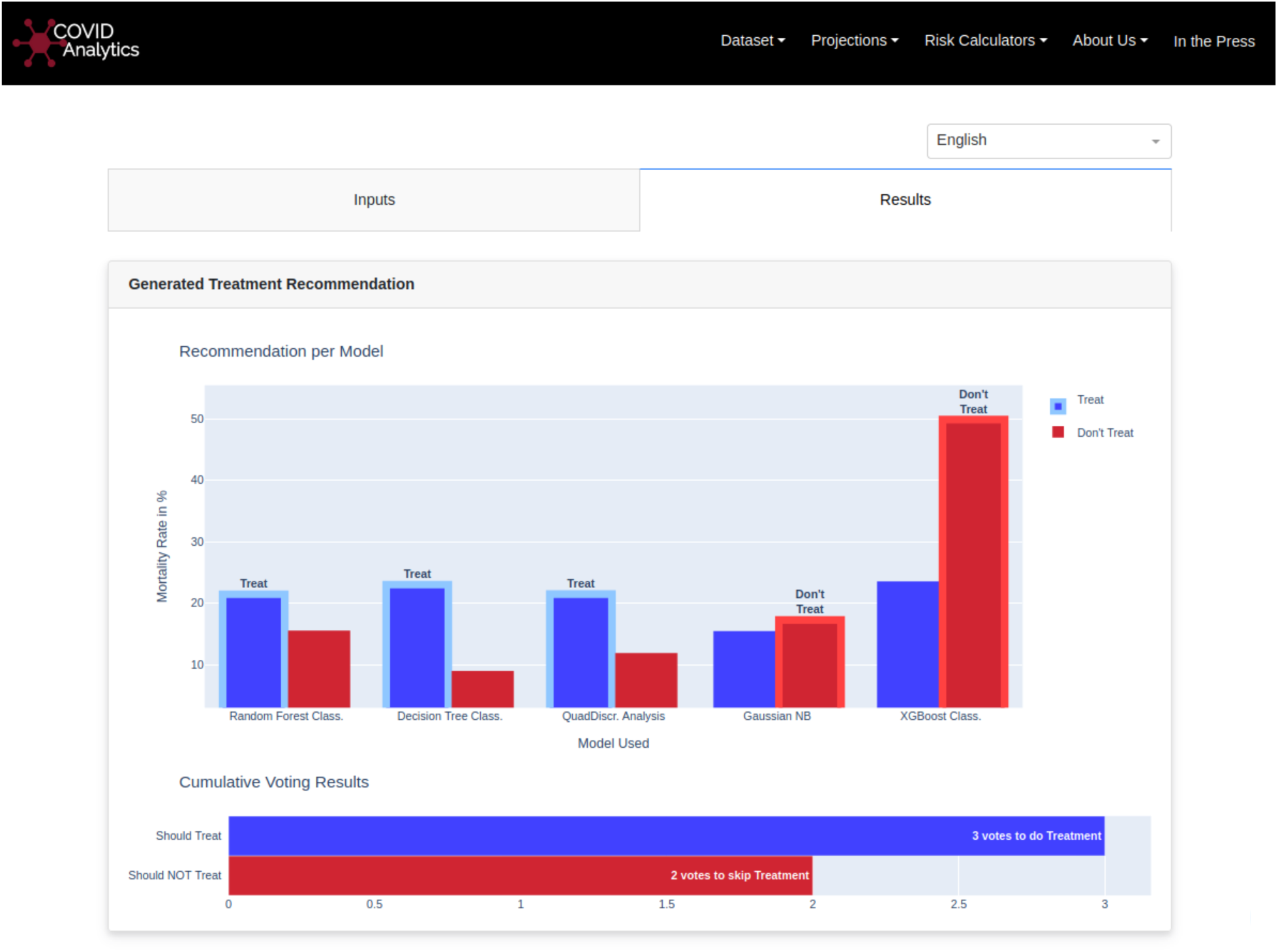
Visualization of the online algorithm user interface.

## 4 Discussion

In this study, we compiled EMR and registry data from five different countries to create predictive and prescriptive models for COVID-19 patients. We demonstrate that accurate analytical models can help physicians assess the potential benefit of ACEI/ARBs for hypertensive patients in practice. To the best of our knowledge, this constitutes the first personalized prescription algorithm for COVID-19 patients that has been validated in an international cohort of patients. We place particular emphasis on essential components of causal inference by applying matching methods to confirm common baseline characteristics between both treatment groups, establishing similar baseline risk between the two populations. We combine six well-established binary classification techniques to predict in-hospital mortality and morbidity. We combine these models on an individual basis, using a voting scheme to assess whether the prescription of ACEI/ARBs can reduce the probability of a hypertensive patient experiencing an adverse event during hospital admission. We apply detailed quantitative evaluation metrics to assess the recommendations’ effectiveness and robustness. We demonstrate through various metrics that the application of our framework can lead to improved patient outcomes relative to the standard of care.

### 4.1 Clinical Insights

Figure 3 offers insight into how our proposed treatment scheme agrees with and differs from what was observed in the data, as measured by the prescription rates. Our algorithm recommends an increase in the overall number of prescriptions of ACEI/ARBs. This includes suggestions to hypertensive patients who were not originally prescribed this line of therapy as well as patients who were already prescribed this class of drugs. A benefit of personalization is also the identification of patients for whom getting this regimen might be detrimental.

Figure 3(a) shows that the algorithm recommends a higher ACEI/ARB prescription rate for patients with heart disease. This could be highly impactful given the prevalence and potential consequences of cardiovascular disease in severe COVID-19 cases [16, 38]. In particular, we note that the algorithm identifies a significantly higher proportion of patients with atrial fibrillation who would benefit from this class of drugs. This finding is in line with the hypothesis that this comorbidity in combination with COVID-19 can lead to severe complications [49], and potentially extends previous findings suggesting a benefit of ACEI/ARBs in relation to atrial fibrillation [26].

The proposed personalized treatment allocation identifies a potential subgroup of hypertensive patients for which ACEI/ARB prescriptions may be detrimental. Figure 3(d) highlights that the prescription rate should be lowered amongst patients with low oxygen based on the mortality/morbidity outcome. Oxygen saturation has been commonly used as a reference metric to potentially identify respiratory complications due to COVID-19. This finding provides an interesting direction for future clinical research.

Our prescription scheme proposes an increase in prescription rates across all age groups, as indicated in Figure 3(e). The algorithm proposes the most significant increase in prescription rates for the youngest cohort below 40 years of age.

Other clinical criteria for ACEI/ARBs cannot be confirmed by our study due to our outcome of interest. For example, ACE inhibitors are known to be risky for women who may become pregnant due to potential birth defects, [44] yet our prescription scheme proposes a higher prescription rate for women (Figure 3(g)). This is not surprising, because such an effect would not be captured in our mortality/morbidity outcome. These external factors demonstrate the need for clinical expertise; while this tool can facilitate treatment decisions, it must be considered in the broader context of a patient’s care.

Finally, we note that our criteria for prescription is an improvement of at least 5.0% in predicted mortality/morbidity during hospitalization. Thus, the reduction does not necessarily imply that ACEI/ARBs are harmful to the remaining patients; the effect could be neutral. The raw predicted probabilities for a patient under each treatment alternative can be assessed in more detail on an individual basis. This can assist clinicians by quantifying the effect of ACEI/ARBs on the specific outcome of interest, mortality/morbidity from COVID-19, which can then be traded off against other external concerns such as chronic hypertension treatment for.

### 4.2 Limitations

There are several limitations to this study. We consider the effect of ACEI/ARBs in isolation, rather than in combination with other treatments. This assumes that the effect of other treatments is independent from the effect of ACEI/ARBs. This is consistent with existing literature, in which COVID-19 treatments have generally been considered separately rather than as combination regimens. However, there is potential to consider treatment strategies more holistically as drug combinations. The methodology presented in this work could be easily extended to the case of multiple treatments: rather than training models for the treated and untreated group, as done here, models could be trained for *N* treatment groups, and the same prescription and voting scheme could be followed to choose between the *N* alternatives. This was impractical in the current study given the scope of the available data, as the sample sizes become much smaller when dividing the population by treatment combination, but this could be considered in future work as larger datasets become available.

All of the data included in the derivation and validation cohorts were collected between February to May 2020. As a result, our investigation carries the limitations associated with the design of observational studies. Moreover, we would like to highlight that the outcome prevalence seems to be dependent on the relative timing of the pandemic curve. Hence, confounding factors such as the degree of congestion in the hospital systems, or changes in the clinical protocols and the use of other drugs might have affected the observed mortality and morbidity rates.

Additionally, there is a tradeoff between obtaining detailed clinical data and curating large datasets. In order to leverage a broad international cohort of patients, we were unable to use granular data that was only available for subgroups of patients. As a result, we used binary indicators with predefined cutoff values for many clinical features. If raw lab readings were available, we could gain further insight into these features and identify data-driven risk cutoffs.

Finally, we note that these results are not causal and do not isolate the effect of ACEI/ARBs in patient outcomes. However, given the time and cost involved in implementing an RCT, we believe that this study adds value. Our study provides insight into potential subpopulations with maximal benefit from ACEI/ARBs that can guide future clinical studies.

## 5 Conclusions

Our approach provides promising evidence for the the benefit of individualizing ACEI/ARBs for hypertensive COVID-19 patients. Using machine learning, we are able to identify patients who would benefit the most by receiving this type of medication. Our framework highlights the potential effect of this class of drugs for hypertensive patients or cases admitted with low oxygen saturation. By personalizing the drug prescription process, the proposed framework improves patient outcomes and avoids unnecessary drug prescriptions that would have limited efficacy. In the future, the algorithm could be integrated in practice into existing EMR systems to generate dynamically personalized treatment recommendations. Our data-driven approach invites further testing using datasets from other hospitals or other types of treatment. Our work is a key step toward a fully patient-centered approach to COVID-19 management and the utilization of existing treatments to reduce its toll on public health.

## Data Availability

The data used in this study are available upon request as they contain potentially identifying or sensitive patient information. This policy follows the restrictions imposed by the the independent healthcare organizations and the Massachusetts Institute of Technology institutional review boards that approved the study. Electronic health record data cannot be shared publicly because they consists of personal information from which it is difficult to guarantee de-identification. As a result, there is a possibility of deductive disclosure of participants and therefore full data access through a public repository is not permitted by the institutions that provided us the data. The data and associated documentation from each collaborating institution can only be made available under a new data sharing agreement with which includes: 1) commitment to using the data only for research purposes and not to identify any individual participant; 2) a commitment to securing the data using appropriate measures, and 3) a commitment to destroy or return the data after analyses are complete. Requests can be made directly at.

## Acknowledgements

The authors wish to thank Travis Ziegler for his help in creating the online interface of the algorithm. We would like to thank the clinical team from Azienda Socio-Sanitaria Territoriale di Cremona (ASST Cremona) and HM Hospitals for collecting their electronic health records, creating a COVID-19 dataset, and providing us with access to it. The authors are also grateful to the HOPE collaboration for compiling a comprehensive registry from multiple countries and centers and sharing it with us.

