## Supplemental Material for "Personalized Prescription of ACEI/ARBs for Hypertensive COVID-19 Patients"

#### 1 Derivation of Comorbidities, Medications, and Lab Values from Patient Data

The dataset we use for modeling is comprised of datasets from several different sources. Data originating from HOPE is sourced from a COVID-19 specific clinical evaluation registry. For each patient included in this registry, the database contains variables for demographics, clinical history of comorbidities and medications in a binary (yes/no) format, hospital admission assessment variables such as maximum temperature and results of laboratory measurements, and a record of comordibities that were developed and medications that were administered during the patient's hospital stay in a binary (yes/no) format. We utilized the HOPE dataset as a template for the format of the final dataset and, consequently, reformatted the other datasets to match. Additional information regarding the HOPE registry can be found here: https://hopeprojectmd.com/en/.

Data from HM Hospitals, ASST Cremona, and Brigham and Women's Hospital (BWH) are sourced from electronic medical record (EMR) systems that collected information on patients throughout their COVID-19 hospitalization, from admission through discharge or death. The EMR data include demographics, vital signs at admission, measurements resulting from laboratory requests during admission, medications administered during admission (identified by their brand name and classification), and records of diagnostic and procedural information coded according to the ICD format which correspond either to pre-admission episodes or inpatient episodes. In order to obtain a cohesive set of comorbidity and medication covariates across the different datasets, we utilized external sources for mapping purposes.

To align how comorbidities were recorded, we made use of Healthcare Cost and Utilization Project (HCUP) tools to map ICD diagnosis codes to comorbidity variables that existed in the HOPE registry data. In Table S1, we list each comorbidity of interest from the HOPE dataset, as well as the corresponding HCUP single-level Clinical Classifications Software (CCS) diagnosis category labels (Agency for Healthcare Research and Quality 2019).

Table S1: Comorbidity variables and corresponding HCUP Category Labels.

| HOPE Registry Comorbidity | HCUP Single-Level CCS Diagnosis Category Labels |
|---|---|
| Diabetes | 49, 50, 186 |
| Hypertension | 98, 99, 183 |
| Dislipidemia | 53 |
| Obesity | 58 |
| Renal Insufficiency | 157, 158 |
| Any Lung Disease | 127, 128, 132, 133 |
| Atrial Fibrillation | 106 |
| Any Heart Disease | 101, 103, 104, 106 |
| Any Cerebrovascular Disease | 109, 111, 112, 113 |
| Connective Disease | 210, 211 |
| Liver Disease | 6, 151 |
| Cancer | 11, 12, 13, 14, 15, 16, 17, 18, 19, 20, 21, 22, 23, 24, 25, 26, 27, 28, 29, 30, 31, 32, 33, 34, 35, 36, 37, 38, 39, 40, 41, 42, 43 |
| Sepsis | 2 |
| Acute Renal Failure | 157 |
| Heart Failure | 108 |
| Embolic Event | 116 |

To standardize the medication variables according to the medication categories recorded in the HOPE registry, we used drug mappings available within the EMR. The BWH data had multiple levels of classification available directly in the EMR which allowed us to group together drugs. The European EMRs recorded the ATC Classification System within the record which allowed us to group individual medications together. The ATC Classification System divides substances into groups (according to the organ or system on which they act and on their therapeutic, pharmacological, and chemical proprieties) through four levels. In Table S2, we list the medication covariates from HOPE and the corresponding ATC codes from the HM Hospitals and ASST Cremona datasets which were used to determine which patients received listed medication. Different levels of ATC classification were used depending on the medication of interest.

Aligning lab values across the different datasets was more straightforward. For continuous lab values, the only required transformations were metric conversions. For binary lab values, the HOPE registry provided cutoff values, and these cutoff values were applied to the other datasets to transform continuous measurements to binary variables.

Table S2: Medication variables and corresponding ATC Codes.

| HOPE Registry Medications | ATC Classification System Codes |
|---|---|
| Corticosteroids | H02 |
| Interferonor | L03AB |
| Tocilizumab | L04AC07 |
| Antibiotics | J01 |
| ACE Inhibitors or ARBs | C09 |
| Cloroquine | P01BA |
| Antiviral | J05AR |
| Anticoagulants | B01AB |

### 2 Missing Data Imputation

Missing values are prevalent, even at small percentages, in the majority of the included variables. Some patients were missing specific elements of the set of collected variables in the HOPE registry. Many EMR observations (HM Foundation, Cremona, and BWH) were incomplete as there was not a concrete list of risk factors to be recorded during data collection. Missing values were imputed using the R implementation of the well-established Multivariate Imputation by Chained Equations (MICE) package (Buuren and Groothuis-Oudshoorn 2010). The use of missing data imputation techniques instead of complete case analysis permits the inclusion of a wider set of features which otherwise would have been omitted in our analysis. MICE initially fills all missing values with an initial random solution. Subsequently, the first variable with at least one missing value, i.e., $p_1$, is regressed on all other variables. All unknown values in $p_1$ are then imputed by simulated draws from the posterior predictive distribution of $p_1$. The same procedure is applied to all other features using the newly imputed values of the previous covariates. To ensure stability, the process is repeated for 10 iterations to produce the final imputed dataset. We excluded all columns whose missing percentage level was higher than 30% at the HOPE dataset. The derivation cohort was imputed independently of the derivation population set to avoid any bias in the resulting data. Tables S3-S4 compare the distribution of the clinical features in the derivation and validation cohorts after data imputation, grouped by data source.

Table S3: Comparison of clinical features in derivation population after data imputation, grouped by data source. Count (proportion) is reported for binary variables. Median (IQR) is presented for continuous features.

| | Hope-Spain | HM-Spain |
|---|---|---|
| Patient Count | 2209 | 497 |
| Age | 69.0 (55.0-78.0) | 74.0 (65.0-83.0) |
| Gender = Male | 1277 (57.8%) | 307 (61.8%) |
| Race = Black | 3 (0.1%) | 0 (0.0%) |
| Race = Caucasian | 1990 (90.1%) | 497 (100.0%) |
| Race = Hispanic | 187 (8.5%) | 0 (0.0%) |
| Race = Asian | 9 (0.4%) | 0 (0.0%) |
| Temperature | 36.8 (36.8-37.5) | 36.6 (36.2-37.2) |
| Creatinine (mg/dL) | 0.9 (0.7-1.1) | 1.0 (0.8-1.3) |
| Sodium (mmol/L) | 138.0 (135.0-140.0) | 136.6 (134.4-139.1) |
| Hemoglobin (g/dL) | 14.0 (12.0-15.0) | 14.0 (12.8-15.0) |
| Leukocytes (1e3/muL) | 6.2 (4.8-8.4) | 6.7 (5.2-9.0) |
| Lymphocytes (1e3/muL) | 1.0 (0.7-1.4) | 1.0 (0.7-1.4) |
| Platelets (1e3/muL) | 193.0 (150.0-251.0) | 207.0 (158.5-258.0) |
| Low Oxygen Saturation | 793 (35.9%) | 166 (33.4%) |
| Low Systolic BP | 239 (10.8%) | 30 (6.0%) |
| Elevated D-Dimer | 1588 (71.9%) | 384 (77.3%) |
| Elevated CRP | 2050 (92.8%) | 459 (92.4%) |
| Elevated Transaminases | 901 (40.8%) | 193 (38.8%) |
| Elevated LDH | 1726 (78.1%) | 330 (66.4%) |
| Diabetes | 678 (30.7%) | 134 (27.0%) |
| Hypertension | 2209 (100.0%) | 497 (100.0%) |
| Dislipidemia | 1162 (52.6%) | 178 (35.8%) |
| Obesity | 681 (30.8%) | 87 (17.5%) |
| Renal Insufficiency | 256 (11.6%) | 20 (4.0%) |
| Lung Disease | 531 (24.0%) | 184 (37.0%) |
| Atrial Fibrillation | 140 (6.3%) | 91 (18.3%) |
| HIV | 6 (0.3%) | 0 (0.0%) |
| Heart Disease | 804 (36.4%) | 159 (32.0%) |
| Cerebrovascular Disease | 284 (12.9%) | 2 (0.4%) |
| Connective Tissue Disease | 79 (3.6%) | 45 (9.1%) |
| Liver Disease | 108 (4.9%) | 33 (6.6%) |
| Cancer | 396 (17.9%) | 69 (13.9%) |
| Corticosteroids | 884 (40.9%) | 227 (45.7%) |
| Interferons | 258 (12.0%) | 26 (5.2%) |
| Tocilizumab | 191 (8.6%) | 81 (16.3%) |
| Antibiotics | 1746 (79.0%) | 463 (93.2%) |
| Death | 455 (20.6%) | 116 (23.3%) |
| Mortality/Morbidity | 723 (32.7%) | 151 (30.4%) |
| Heart Failure | 141 (6.4%) | 24 (4.8%) |
| Acute Renal Failure | 376 (17.0%) | 53 (10.7%) |
| Sepsis | 251 (11.4%) | 5 (1.0%) |
| Embolic Event | 39 (1.8%) | 0 (0.0%) |
| Mechanical Ventilation | 125 (5.7%) | 10 (2.0%) |

Table S4: Comparison of clinical features in validation population after data imputation, grouped by data source. Count (proportion) is reported for binary variables. Median (IQR) is presented for continuous features.

| | Cremona-Italy | Hope-Other | MGB-USA |
|---|---|---|---|
| Patient Count | 249 | 375 | 177 |
| Age | 72.0 (63.0-80.0) | 60.0 (50.0-74.0) | 74.0 (63.0-85.0) |
| Gender = Male | 161 (64.7%) | 257 (68.5%) | 93 (52.5%) |
| Race = Black | 0 (0.0%) | 0 (0.0%) | 48 (27.1%) |
| Race = Caucasian | 249 (100.0%) | 262 (69.9%) | 95 (53.7%) |
| Race = Hispanic | 0 (0.0%) | 110 (29.3%) | 30 (16.9%) |
| Race = Asian | 0 (0.0%) | 3 (0.8%) | 3 (1.7%) |
| Temperature | 37.3 (36.6-37.8) | 37.0 (36.8-37.7) | 37.2 (36.7-37.8) |
| Creatinine (mg/dL) | 1.1 (0.9-1.4) | 0.9 (0.8-1.2) | 1.1 (0.8-1.5) |
| Sodium (mmol/L) | 138.0 (136.0-140.0) | 137.0 (134.0-139.0) | 139.0 (137.0-141.0) |
| Hemoglobin (g/dL) | 13.6 (12.3-14.9) | 14.0 (12.0-15.0) | 11.6 (10.3-12.8) |
| Leukocytes (1e3/muL) | 7.2 (5.3-9.8) | 6.8 (5.0-9.8) | 6.8 (5.1-8.8) |
| Lymphocytes (1e3/muL) | 1.0 (0.7-1.4) | 1.0 (0.8-1.4) | 1.0 (0.8-1.4) |
| Platelets (1e3/muL) | 189.0 (148.0-262.0) | 209.0 (157.5-291.0) | 200.0 (144.0-264.6) |
| Low Oxygen Saturation | 105 (42.2%) | 172 (45.9%) | 19 (10.7%) |
| Low Systolic BP | 9 (3.6%) | 28 (7.5%) | 8 (4.5%) |
| Elevated D-Dimer | 236 (94.8%) | 239 (63.7%) | 168 (94.9%) |
| Elevated CRP | 115 (46.2%) | 335 (89.3%) | 172 (97.2%) |
| Elevated Transaminases | 133 (53.4%) | 142 (37.9%) | 89 (50.3%) |
| Elevated LDH | 73 (29.3%) | 246 (65.6%) | 32 (18.1%) |
| Diabetes | 24 (9.6%) | 92 (24.5%) | 102 (57.6%) |
| Hypertension | 249 (100.0%) | 375 (100.0%) | 177 (100.0%) |
| Dislipidemia | 5 (2.0%) | 157 (41.9%) | 109 (61.6%) |
| Obesity | 5 (2.0%) | 116 (30.9%) | 62 (35.0%) |
| Renal Insufficiency | 0 (0.0%) | 36 (9.6%) | 47 (26.6%) |
| Lung Disease | 4 (1.6%) | 90 (24.0%) | 73 (41.2%) |
| Atrial Fibrillation | 9 (3.6%) | 9 (2.4%) | 42 (23.7%) |
| HIV | 0 (0.0%) | 1 (0.3%) | 0 (0.0%) |
| Heart Disease | 22 (8.8%) | 134 (35.7%) | 69 (39.0%) |
| Cerebrovascular Disease | 0 (0.0%) | 36 (9.6%) | 34 (19.2%) |
| Connective Tissue Disease | 1 (0.4%) | 5 (1.3%) | 46 (26.0%) |
| Liver Disease | 0 (0.0%) | 11 (2.9%) | 23 (13.0%) |
| Cancer | 0 (0.0%) | 55 (14.7%) | 41 (23.2%) |
| Corticosteroids | 147 (59.0%) | 191 (52.8%) | 25 (14.1%) |
| Interferons | 8 (3.2%) | 89 (24.7%) | 0 (0.0%) |
| Tocilizumab | 6 (2.4%) | 34 (9.1%) | 0 (0.0%) |
| Antibiotics | 186 (74.7%) | 318 (84.8%) | 116 (65.5%) |
| Death | 77 (30.9%) | 90 (24.0%) | 39 (22.0%) |
| Mortality/Morbidity | 78 (31.3%) | 126 (33.6%) | 57 (32.2%) |
| Heart Failure | 1 (0.4%) | 26 (6.9%) | 7 (4.0%) |
| Acute Renal Failure | 0 (0.0%) | 48 (12.8%) | 14 (7.9%) |
| Sepsis | 0 (0.0%) | 30 (8.0%) | 4 (2.3%) |
| Embolic Event | 0 (0.0%) | 11 (2.9%) | 0 (0.0%) |
| Mechanical Ventilation | 4 (1.6%) | 63 (16.8%) | – |

### 3 Covariate Matching

In this section, we present a detailed descriptive summary of the derivation population broken down into the treatment and control group. We distinguish the distribution of features before and after the matching process to demonstrate the effect of the covariate balancing step.

Table S5: Comparison of clinical features in derivation population between treatment groups, before and after matching procedure. Count (proportion) is reported for binary variables. Median (IQR) is presented for continuous features.

| | Pre-Match | | Post-Match | |
|---|---|---|---|---|
| | ACEI/ARBs | No ACEI/ARBs | ACEI/ARBs | No ACEI/ARBs |
| Patient Count | 1043 | 1663 | 960 | 960 |
| Age | 70.0 (57.0-79.0) | 70.0 (56.5-79.0) | 71.0 (57.0-80.0) | 71.0 (58.0-79.0) |
| Gender = Male | 599 (57.4%) | 985 (59.2%) | 557 (58.0%) | 557 (58.0%) |
| Race = Black | 2 (0.2%) | 1 (0.1%) | 2 (0.2%) | 1 (0.1%) |
| Race = Caucasian | 955 (91.6%) | 1532 (92.1%) | 885 (92.2%) | 890 (92.7%) |
| Race = Hispanic | 78 (7.5%) | 109 (6.6%) | 66 (6.9%) | 56 (5.8%) |
| Race = Asian | 3 (0.3%) | 6 (0.4%) | 3 (0.3%) | 3 (0.3%) |
| Temperature | 36.8 (36.5-37.5) | 36.8 (36.6-37.4) | 36.8 (36.6-37.5) | 36.8 (36.6-37.4) |
| Creatinine (mg/dL) | 0.9 (0.7-1.2) | 0.9 (0.7-1.2) | 0.9 (0.7-1.2) | 0.9 (0.7-1.2) |
| Sodium (mmol/L) | 138.0 (135.0-140.0) | 138.0 (135.0-140.0) | 138.0 (135.0-140.0) | 138.0 (135.0-140.0) |
| Hemoglobin (g/dL) | 14.0 (12.5-15.0) | 14.0 (12.9-15.0) | 14.0 (12.3-15.0) | 14.0 (12.2-15.0) |
| Leukocytes (1e3/muL) | 6.4 (4.9-8.5) | 6.3 (4.8-8.4) | 6.4 (4.8-8.5) | 6.4 (4.9-8.5) |
| Lymphocytes (1e3/muL) | 1.0 (0.7-1.4) | 1.0 (0.7-1.4) | 1.0 (0.7-1.4) | 1.0 (0.7-1.4) |
| Platelets (1e3/muL) | 194.0 (150.0-255.0) | 195.0 (152.0-250.0) | 193.0 (151.0-254.0) | 195.0 (152.0-254.2) |
| Low Oxygen Saturation | 345 (33.1%) | 614 (36.9%) | 325 (33.9%) | 325 (33.9%) |
| Low Systolic BP | 78 (7.5%) | 191 (11.5%) | 77 (8.0%) | 77 (8.0%) |
| Elevated D-Dimer | 728 (69.8%) | 1244 (74.8%) | 682 (71.0%) | 682 (71.0%) |
| Elevated CRP | 945 (90.6%) | 1564 (94.0%) | 888 (92.5%) | 888 (92.5%) |
| Elevated Transaminases | 406 (38.9%) | 688 (41.4%) | 390 (40.6%) | 390 (40.6%) |
| Elevated LDH | 758 (72.7%) | 1298 (78.1%) | 711 (74.1%) | 711 (74.1%) |
| Diabetes | 315 (30.2%) | 497 (29.9%) | 297 (30.9%) | 286 (29.8%) |
| Hypertension | 1043 (100.0%) | 1663 (100.0%) | 960 (100.0%) | 960 (100.0%) |
| Dislipidemia | 517 (49.6%) | 823 (49.5%) | 475 (49.5%) | 475 (49.5%) |
| Obesity | 310 (29.7%) | 458 (27.5%) | 282 (29.4%) | 282 (29.4%) |
| Renal Insufficiency | 84 (8.1%) | 192 (11.5%) | 82 (8.5%) | 82 (8.5%) |
| Lung Disease | 285 (27.3%) | 430 (25.9%) | 257 (26.8%) | 257 (26.8%) |
| Atrial Fibrillation | 70 (6.7%) | 161 (9.7%) | 63 (6.6%) | 103 (10.7%) |
| HIV | 2 (0.2%) | 4 (0.2%) | 2 (0.2%) | 2 (0.2%) |
| Heart Disease | 360 (34.5%) | 603 (36.3%) | 326 (34.0%) | 326 (34.0%) |
| Cerebrovascular Disease | 96 (9.2%) | 190 (11.4%) | 94 (9.8%) | 94 (9.8%) |
| Connective Tissue Disease | 52 (5.0%) | 72 (4.3%) | 48 (5.0%) | 39 (4.1%) |
| Liver Disease | 66 (6.3%) | 75 (4.5%) | 60 (6.2%) | 46 (4.8%) |
| Cancer | 155 (14.9%) | 310 (18.6%) | 145 (15.1%) | 165 (17.2%) |
| Corticosteroids | 387 (38.0%) | 724 (44.1%) | 371 (38.6%) | 371 (38.6%) |
| Interferons | 108 (10.6%) | 176 (10.8%) | 102 (10.6%) | 102 (10.6%) |
| Tocilizumab | 96 (9.2%) | 176 (10.6%) | 93 (9.7%) | 105 (10.9%) |
| Antibiotics | 842 (80.7%) | 1367 (82.2%) | 778 (81.0%) | 795 (82.8%) |
| Death | 219 (21.0%) | 352 (21.2%) | 214 (22.3%) | 186 (19.4%) |
| Mortality/Morbidity | 329 (31.5%) | 545 (32.8%) | 314 (32.7%) | 301 (31.4%) |
| Heart Failure | 73 (7.0%) | 92 (5.5%) | 69 (7.2%) | 58 (6.0%) |
| Acute Renal Failure | 163 (15.6%) | 266 (16.0%) | 157 (16.4%) | 146 (15.2%) |
| Sepsis | 108 (10.4%) | 148 (8.9%) | 105 (10.9%) | 83 (8.6%) |
| Embolic Event | 14 (1.3%) | 25 (1.5%) | 14 (1.5%) | 10 (1.0%) |
| Mechanical Ventilation | 53 (5.1%) | 82 (4.9%) | 52 (5.4%) | 46 (4.8%) |

### Predictive Modeling Details

In this section, we provide additional details regarding the parameter tuning process of the binary classification algorithms and their discrimination performance across all data sets considered.

*Parameter Tuning*

One of the key steps of a machine learning pipeline consists in training the model that is used to predict the desired outcome. This training procedure involves a trade off between the performance of the model on the training population, and its predicting power on an external (unseen) population. One of the most common errors in the training step is overfitting: the model gets excessive exposure to the training population and fails to learn the predictive signal in the data, so that it achieves a strong performance in sample, but it is not able to accurately predict the outcome on unseen data. This can be mitigated by tuning the model parameters using a validation procedure, which finds the optimal parameters for a given algorithm by making sure that the resultant models are able to achieve high accuracy on new data points. First, we define the search space for the parameters of each model and then we use the Optuna framework (Akiba et al. 2019) to maximize the objective function defined as the 20-fold Cross Validation Area Under the Curve ($AUC$). All the procedure is pursued by leveraging different cores over 300 iterations.

*Methods Comparison*

A comparison of the seven machine learning methods used to predict the binary outcome is presented in Table S6. In all cases, we formulate a binary classification problem to predict mortality/morbidity as the endpoint of a patient's hospitalization. Predictive models are trained using Logistic Regression (LR), Random Forest (RF), Classification And Regression Trees (CART), Optimal Classification Trees (OCT), Gradient Boosted Trees (XGBoost), Quadratic Discriminant Analysis (QDA), and Gaussian Naive Bayes (GB); all methods are implemented in Scikit-learn (Pedregosa et al. 2011) other than XGBoost which was trained using the XGBoost package (Chen and Guestrin 2016). Logistic Regression assumes an additive relationship between risk factors resulting in a significantly lower performance compared to the other methods. Thus, it was excluded from the prescriptive component of the proposed framework. Table S6 compares the discrimination performance (AUC) of the trained binary classification models on training, testing, and external validation sets broken down into the treatment and control group.

Table S6: Predictive model AUC performance results with Logistic Regression included as an additional machine learning method.

| | Training Data | | Testing Data | | Validation Data | |
|---|---|---|---|---|---|---|
| | ACEI/ARBs | No ACEI/ARBs | ACEI/ARBs | No ACEI/ARBs | ACEI/ARBs | No ACEI/ARBs |
| **LR** | 0.797 | 0.773 | 0.786 | 0.747 | 0.660 | 0.700 |
| **RF** | 0.886 | 0.862 | 0.834 | 0.814 | 0.770 | 0.761 |
| **CART** | 0.843 | 0.827 | 0.812 | 0.790 | 0.762 | 0.714 |
| **OCT** | 0.879 | 0.829 | 0.797 | 0.777 | 0.737 | 0.699 |
| **XGBOOST** | 0.909 | 0.927 | 0.819 | 0.802 | 0.768 | 0.743 |
| **QDA** | 0.883 | 0.870 | 0.827 | 0.813 | 0.718 | 0.736 |
| **GB** | 0.824 | 0.813 | 0.826 | 0.805 | 0.710 | 0.734 |
| **Average AUC** | 0.860 | 0.843 | 0.814 | 0.793 | 0.732 | 0.727 |

*Feature Importance for Predictive Models*

The most important model features, as measured by the SHAP value and averaged across methods, are presented in Table S7. Creatinine, low oxygen saturation, white blood cell count, hemoglobin, and age are the most important features in both the models with ACEI/ARBs and without ACEI/ARBs. These observations are consistent with Table 4 in the main manuscript, which measures feature importance by average variable rank across the methods.

Table S7: Summary of variable importance for each model by mean absolute SHAP value, where higher values indicate more importance. OCT is not implemented natively in Python and cannot be evaluated using the SHAP methodology.

| | Algorithm | CART | GB | OCT | QDA | RF | XGBOOST | Average |
|---|---|---|---|---|---|---|---|---|
| ACEI/ARBs | Creatinine | 0.143 | – | – | 0.072 | 0.052 | 0.001 | 0.067 |
| | White Blood Cell Count | – | 0.089 | – | 0.109 | 0.020 | 0.000 | 0.055 |
| | Low Oxygen Saturation | 0.105 | – | – | 0.056 | 0.046 | 0.001 | 0.052 |
| | Hemoglobin | – | 0.050 | – | – | – | – | 0.050 |
| | Age | 0.047 | 0.094 | – | 0.061 | 0.040 | 0.001 | 0.048 |
| | Platelets | – | 0.039 | – | 0.056 | – | – | 0.048 |
| | Lymphocytes | – | 0.037 | – | – | 0.018 | 0.000 | 0.019 |
| No ACEI/ARBs | White Blood Cell Count | – | 0.067 | – | 0.092 | – | – | 0.079 |
| | Creatinine | 0.154 | 0.037 | – | 0.064 | 0.027 | 0.005 | 0.058 |
| | Age | 0.056 | 0.095 | – | 0.080 | 0.028 | 0.004 | 0.053 |
| | Low Oxygen Saturation | 0.084 | – | – | 0.043 | 0.023 | 0.004 | 0.039 |
| | Hemoglobin | – | 0.052 | – | 0.060 | 0.011 | 0.002 | 0.031 |
| | Blood Sodium | – | 0.037 | – | – | – | 0.001 | 0.019 |
| | Lymphocytes | 0.016 | – | – | – | 0.013 | – | 0.014 |
| | Temperature | 0.007 | – | – | – | – | – | 0.007 |

### 4 Prescriptive Framework Details

In this section, we delve into the results of the prescriptive framework. We compare the prescription evaluation metrics for different improvement thresholds and also examine each algorithm's prescription frequency and agreement with the proposed voting scheme.

*Choice of Improvement Threshold*

Table S8 outlines the match rate, prescription count, average AUC, prescription effectiveness (PE), calibrated prescription effectiveness (CPE), and prescription robustness (PR) for all improvement thresholds included in our analysis. The threshold of 5% improvement was selected for its balance of reducing prescriptions while maintaining competitive performance in the other metrics, particularly PR.

Table S8: Comparison of key metrics on the test set, assessed at different improvement thresholds.

| | Match Rate | Presc. Count | Avg. AUC | PE | CPE | PR (Low) | PR (High) |
|---|---|---|---|---|---|---|---|
| **0.00** | 0.514 | 160 | 0.800 | 0.006 | 0.005 | -0.007 | -0.035 |
| **0.01** | 0.507 | 154 | 0.806 | 0.009 | 0.008 | -0.007 | -0.034 |
| **0.02** | 0.517 | 141 | 0.808 | 0.002 | 0.001 | -0.007 | -0.034 |
| **0.05** | 0.514 | 124 | 0.804 | 0.008 | 0.007 | -0.008 | -0.033 |
| **0.10** | 0.514 | 86 | 0.829 | 0.004 | 0.003 | -0.007 | -0.030 |

*Comparison of Methods under the Voting Scheme*

Table S9 indicates how frequently each individual method would prescribe ACEI/ARBs, given an improvement threshold of 5%, as well as how often the method's prescription agrees with the final treatment prescription based on the voting scheme.

Table S9: Breakdown of individual method prescription frequency and agreement with the final voting prescription scheme.

| | Prescription Count | Agreement with Prescription |
|---|---|---|
| **RF** | 158 | 78.5% |
| **CART** | 93 | 67.7% |
| **OCT** | 148 | 64.6% |
| **XGBOOST** | 185 | 74.7% |
| **QDA** | 126 | 68.8% |
| **GB** | 124 | 67.4% |
| **Prescription** | 124 | 100.0% |
